# Analysis of the Health Economics Portfolio Funded by the National Institutes of Health in Response to Published Guidance

**DOI:** 10.1101/2023.03.28.23287876

**Authors:** Caitlin E. Burgdorf, William T. Riley, Farheen Akbar, Abbey D. Zuehlke, Katrina Bibb, Heather D. Eshleman, Ami Shah, Deshiree Belis, Michael Spittel, Paula Fearon

**Affiliations:** Office of Behavioral and Social Sciences Research, National Institutes of Health, Bethesda, MD, 20892; Lexical Intelligence, Rockville, MD 20851

**Keywords:** Health economics, National Institutes of Health, application rates, funding rates, Interrupted Time Series Analysis

## Abstract

Health economics is one of the interdisciplinary sciences that the National Institutes of Health (NIH) supports in order to pursue its overall mission to improve health. In 2015, NIH guidance was published to clarify the type of health economics research that NIH would continue to fund. This analysis aimed to determine if there were changes in the number of health economics applications received and funded by NIH after the release of the guidance. Health economics applications submitted to NIH both before and after the guidance was released were identified using a machine learning approach with input from subject matter experts. Application and funding trends were examined by fiscal year, method of application (solicited vs. unsolicited), and activity code. This study found that application and funding rates of health economics research was decreasing prior to guidance. Following publication of this guidance, the application and funding rate of health economics applications increased.

## 1. Introduction

Health economics is defined by the NIH as “the study of how scarce resources are allocated among alternative uses for the care of sickness and the promotion, maintenance, and improvement of health, including lifestyle patterns and behavioral health” (National Institutes of Health, 2015). It includes the study of “how health care and health-related services, their costs, benefits, and consequences, and health itself are distributed among individuals and groups in society by measuring efficiency, effectiveness, value, and impact of health services/resources between individuals, healthcare providers, and clinical settings” (National Institutes of Health, 2015). Health economics research offers rigorous approaches that can help NIH pursue its overall mission to improve health and is an integral part of the interdisciplinary sciences that NIH supports.

To emphasize the types of health economics research that are relevant to the NIH mission, NIH published guidance NOT-OD-16-025 to the extramural community in 2015 (National Institutes of Health, 2015). This guidance specified research areas that were 1) high priority across NIH, 2) priorities that were Institute- and Center (IC)- specific, and 3) those that were not in alignment with the NIH mission. High-priority research included health economics studies designed to understand how innovations in treatment, diagnosis, prevention, and implementation strategies can be deployed most effectively to improve health and well-being as well as research aimed at designing better interventions with these insights. IC-specific priorities included research specific to an IC’s mission utilizing economic methodologies such as assessments of alternative models of delivering IC-mission-relevant care. Assessments focused on outcomes related to health, including health-related behaviors and healthcare utilization. Areas outside of NIH’s mission included studies that were minimally related to specific health outcomes at the population or individual level.

While the goal of the guidance was to communicate that NIH will continue to accept applications involving health economics research with a focus on improving health outcomes and health-related behaviors, the research community demonstrated mixed responses on the perceived influence of this guidance on funding levels within the field (Consortium of Social Science Associations, 2015; Wolinetz, 2015). This study aimed to determine if this guidance had an impact on the number of health economics applications being submitted to the NIH (application rate) or the amount of funding provided to health economics compared to all NIH funding (funding rate). NIH application and funding patterns for research in the field of health economics were analyzed over time, with an emphasis on determining the impact of the 2015 guidance publication on these trends.

## 2. Methods

### 2.1 Identification of the NIH health economics research portfolio

A machine learning (ML) model was created to identify the NIH health economics research portfolio using positive and negative training data. The positive training set was created by combining projects curated by subject matter experts in a 2015 internal analysis (n=198) with health economics research projects identified using keyword searches (n=36) for a total of 234 projects. The negative training set was a subset of research project grant (RPG) awards classified by Research, Condition, and Disease Category (RCDC) as ‘Translational Research’ that were funded by the NIH from FY2010 - FY2020 (n = 3,898).

The machine learning model analyzed features from the text of the title, abstract, specific aims, and research plan of each grant application. Standard text tokenization of the fields was applied including stemming, ignoring case, removing stop words, and collapsing character codes. In addition to using free text, the model also used a controlled vocabulary of health economics terms that was compiled from The British Medical Journal (BMJ Publishing Group Limited, 2021), National Library of Medicine (National Library of Medicine 2021), Centers for Disease Control and Prevention (Centers for Disease Control and Prevention 2021), and the University of York (York Health Economics Consortium, 2021). Gradient Tree Boost and Liblinear algorithms were combined into a single measure using a neural network known as an ensemble algorithm. The model was evaluated for precision (specificity), recall (sensitivity), and F1 score (harmonic mean of precision and recall).

This final ML algorithm was applied to all competing applications (both new applications and renewal applications) for research project grants received by NIH between FY2010 and FY2020 (n=623,676), obtained through the Information for Management Planning Analysis and Coordination (IMPAC) II data system. Applications that were reviewed in council year 2009 or included animal subjects were removed. Each application was given a discrete prediction score of 1 or 0. A prediction score of 1 was assigned if it was predicted to be health economics while a prediction score of 0 was assigned if it was predicted not to be health economics. Each application was also given a confidence score ranging between 0.5 and 1. The higher the confidence scores, the higher likelihood that the prediction was accurate, such that a prediction score of 1 with a confidence score of 0.95 is very likely to be health economics, and conversely a prediction score of 0 with a confidence score of 0.95 is not. A cutoff confidence score of 0.999 was used to capture the health economics portfolio. Subject matter experts validated a sample of results (n=50 applications) for accuracy.

### 2.2 Analysis of trends in health economics research funding

Competing RPG applications that were identified using ML to be health economics (n = 23,362), both awarded (n = 3,661) and not awarded (n=19,701), were compared to competing RPG applications that were not related to health economics (n = 600,314), both awarded (n = 102,649) and not awarded (n = 497,665). Applications and awards within the health economics and non-health economics datasets were analyzed for the number of applications, awards, NIH-designated grant mechanisms and award rates before and after publication of the guidance. As fiscal year and calendar year dates do not align, applications with council review date in January 2016 or before were coded as “pre-guidance” and applications with council review date after January 2016 were coded as “post-guidance”.

An Interrupted Time Series Analysis (ITSA) was used to determine the impact of the release of the 2015 guidance on application and funding rates; an ITSA can evaluate time-dependent outcomes in response to an event by measuring the same outcome parameter at sequential times before and after the event. ITSA was selected as it is the study design of choice for evaluating the effectiveness of interventions with a clearly defined implementation date (Hudson et al. 2019; Bernal et al. 2017). The main utility of the ITSA is the ability to determine whether the pattern in the data observed after the intervention is the same as the pattern pre-intervention.

When analyzing the data through ITSA, all regression models used the following formula: Y_t_ = β_0_ + β_1_T + β_2_D + β_3_P + Ɛ, where:

• Outcome (Y_t_) (application or funding rate) = baseline (β_0_) + time since the start of observations (β_1_T) + treatment/intervention (pre or post) (β_2_D) + time since intervention (β_3_P) + random error (Ɛ)
• Time (T): the time elapsed since the start of the observation period (in this case, council rounds), in frequency terms with which observations are taken/utilized.
• Intervention (D): a dummy variable, described in the data as the “treatment variable,” was used to indicate the pre-guidance period or post-guidance time periods.
  ○ Applications with council review date in January 2016 or before were coded as pre-guidance =0
  ○ Applications with council review date post January 2016 were coded as post-guidance = 1, starting at time=30 (30^th^ council round)
• Time since intervention (P): the time elapsed since the intervention, in frequency terms, with which observations are taken/utilized (council year and month, as seen above). Time in the pre-intervention period (P) was equal to 0, time since intervention (P) started at P = 1 and incrementally increased for each time point (T) following the intervention.
  ○ Discrete variables: 0 (pre-intervention; 1-44 incremental (post-intervention)
  ○ Treatment began at the Time (T) = 30 (January 2016)
• Outcome (Y_t_): the outcome variable (at time t), is the aggregated number of health economics applications or aggregated amount of health economics funding, shown as a rate of the total NIH portfolio; see below for specifics on each outcome rate.
  ○ Application outcome: The application rate for each council round was calculated for the application outcome variable using all applications NIH received during FY2010- FY2020 to calculate application rates at different points in time based on year and council round. The recorded rate was calculated as a rate of health economics applications to non-health economics applications: (HealthEconomicsCountofApplications/Non-HealthEconomicsCountofApplications*1000)
  ○ Total cost outcome: health economics funding rates for each council round were calculated using total costs for health economics awards and total costs for non-health economics awards (i.e., at council round August 2015, the number of dollars for both health economics and NIH were recorded to compute the rate: ((HealthEconomicsSumofTotalCost/Non-HealthEconomicsSumofAwdTotal)*1000)

In addition to assessing the health economics portfolio and non-health economics portfolio, applications and awards were further split into two datasets: solicited and unsolicited. The “solicited” dataset contained applications and subsequent awards that were submitted in response to specific funding announcement. The “unsolicited” dataset contained those that were submitted to a general parent announcement or initiated by the investigator.

## 3. Results

### 3.1 Machine learning approach

The refined ML model that captured the NIH health economics portfolio had an F1 score of >0.93. The threshold confidence score for health economics applications was set at 0.999, a value which was determined by analyzing the projects identified as health economics around this threshold. Due to curating projects around the threshold, the algorithm was thought to have captured health economics projects with high confidence, especially since it was applied uniformly to all applications. However, it is possible with an approach such as this that some non-health economics projects are included in the dataset and that some health economics projects were not included in the dataset. All applications receiving a confidence score less than 0.999 were deemed as non-health economics. The number of applications with each category prediction is shown in Table 1.

**Table 1.** Summary of ML-predicted projects.

| Category Predictions | Total Applications | Non-awarded Applications | Awarded Applications |
| --- | --- | --- | --- |
| Category prediction score= 1 | 43,606 | 37,016 | 6,590 |
| Category prediction= 1, Confidence score $\geq 0.999$ (Health economics) | 23,362 | 19,701 | 3,661 |
| Category prediction 0 (Non-health economics) | 580,070 | 480,350 | 99,720 |

### 3.2 Health economics portfolio overview

#### 3.2.1 Application and award trends by fiscal year

Number of applications and awards for health economics research were compared with applications and awards that were not related to health economics in the pre-guidance period (FY2010- FY2015) as well as the post-guidance period (FY2016- FY2020) (Table 2). In the pre-guidance period, NIH received an average of 2,155 health economics applications per fiscal year, which decreased (not significantly) to 2,086 applications post-guidance. Despite the trending decrease in application numbers in the post-guidance period, the average health economics award count showed a not significant increase from an average of 317 awards per year to an average of 352 awards per year. These trends resulted in a higher award rate (from 14.7% to 16.9%) post-guidance. For applications that were not related to health economics, there was an increase in the number of applications (from 51,127 to 58,710 yearly average), number of awarded applications (from 8,397 to 10,453 yearly average), and the award rate (16.4% to 17.8%). Award rates by fiscal year are detailed in the appendix (Table A1, Figure A1).

**Table 2.** HE and non-HE applications. Yearly average, total counts, and award rates of health economics (HE) applications and applications that were not related to health economics (non-HE) before and following publication of the HE guidance.

| **Examination Period** | **Total HE Applications** | **HE Non-Awarded Applications** | **HE Awarded Applications** | **Award Rate** |
| --- | --- | --- | --- | --- |
| Pre-guidance | 2,155 (yearly average)<br>12,932 (total) | 1,839 (yearly average)<br>11,031 (total) | 317 (yearly average)<br>1,901 (total) | 14.7% |
| Post-guidance | 2,086 (yearly average)<br>10,430 (total) | 1,734 (yearly average)<br>8,670 (total) | 352 (yearly average)<br>1,760 (total) | 16.9% |
| **Examination Period** | **Total Non-HE Applications** | **Non-HE Non-Awarded Applications** | **Non-HE Awarded Applications** | **Award Rate** |
| Pre-guidance | 51,127 (yearly average)<br>306,763 (total) | 42,730 (yearly average)<br>256,381 (total) | 8,397 (yearly average)<br>50,382 (total) | 16.4% |
| Post-guidance | 58,710 (yearly average)<br>293,551 (total) | 48,257 (yearly average)<br>241,284 (total) | 10,453 (yearly average)<br>52,267 (total) | 17.8% |

#### 3.2.2 Solicited vs. unsolicited application and award trends

NIH also releases Funding Opportunity Announcements (FOAs) to solicit applications that align with NIH goals and objectives in addition to accepting investigator-initiated applications. For research that was not related to health economics, 39% of applications and 36% of awards were solicited from FY2010 - FY2020 (Figure 1). However, for research that was related to health economics, 57% of applications and 56% of awards were solicited (Figure 1). This pattern remained consistent across all fiscal years for solicited application rates and respective award rates (see appendix Figure A2 and A3).

**Figure 1.**
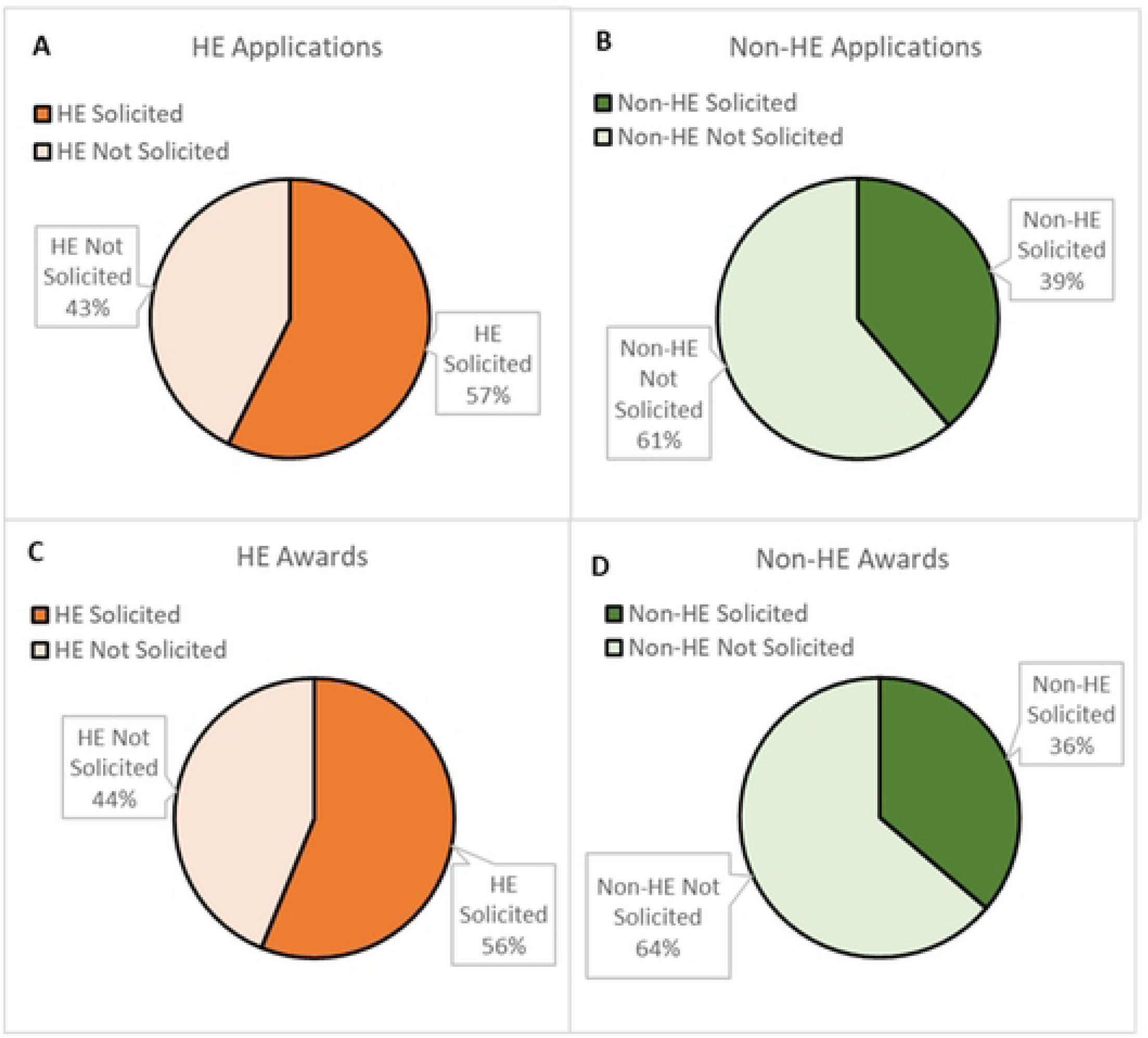
HE and non-HE applications and awards by method of application (solicited or unsolicited) from FY2010- FY2020. Competing applications and awards for HE and non-HE research were determined for FY2010- FY2020 and then broken out by whether they were solicited or unsolicited. [A] HE applications from FY2010- FY2020. [B] Non-HE applications from FY2010- FY2020. [C] HE awards from FY2010- FY2020. [D] Non-HE awards from FY2010- FY2020.

#### 3.2.3 Application and Award Trends by Activity Code

NIH utilizes a number of grant mechanisms for RPGs that vary in the length and amount of funding, identified by activity codes (e.g. R01, R21, R03). The health economics and non-health economics datasets were analyzed to see if variation existed in the distribution of activity codes for health economics research and the rest of the NIH portfolio (Figure 2). Applications and awards for health economics and non-health economics research from FY2010- FY2020 had similar activity codes with R01s making up the majority of applications and awards for health economics research and non-health economics research, followed by R21s and R03s. Activity code types overlapped in all of the top five categories by application counts and four of the top five in award counts.

**Figure 2.**
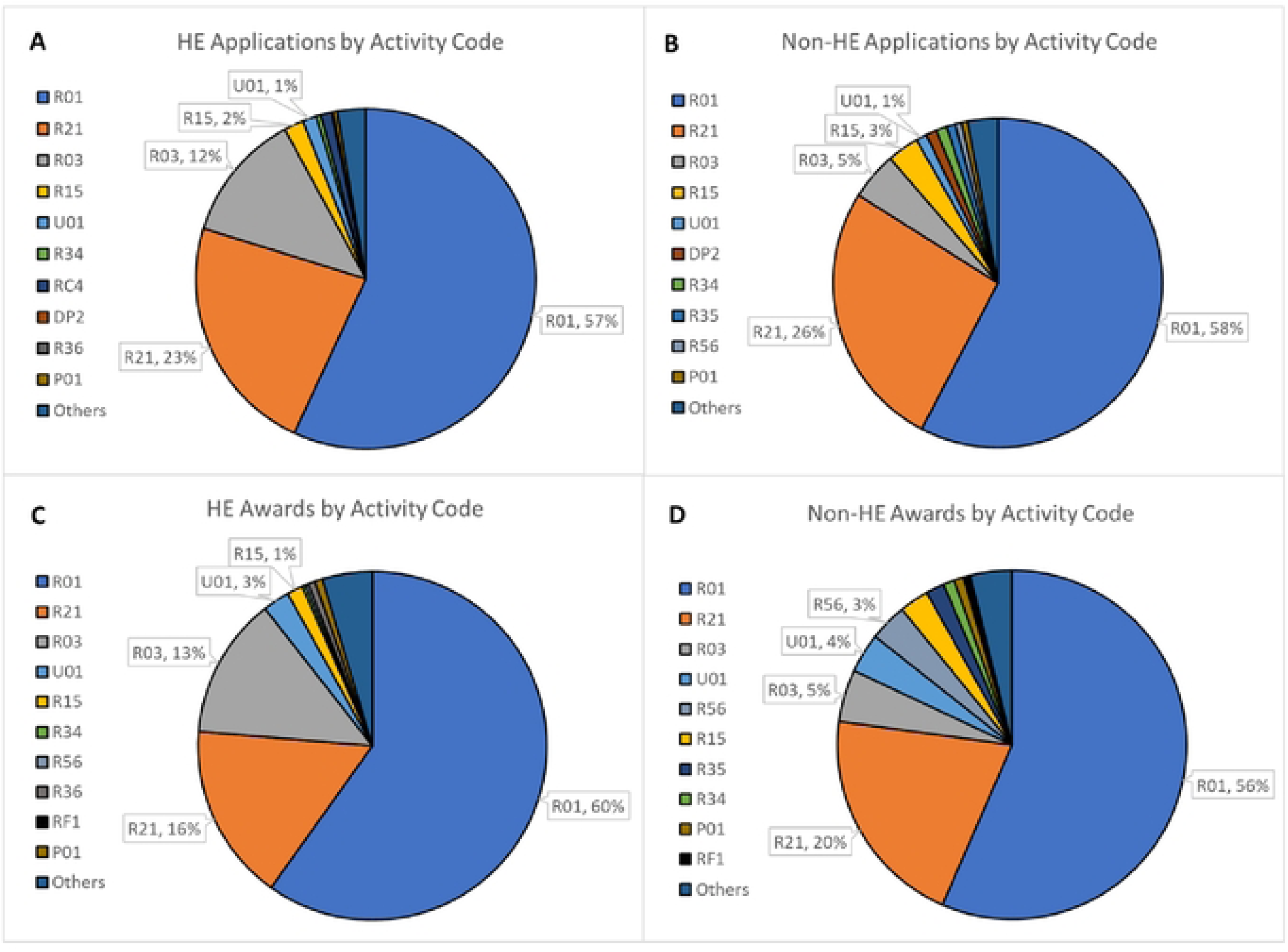
HE and non-HE applications and awards by activity code from FY2010- FY 2020. Competing applications and awards for HE and non-HE research were determined for FY2010- FY2020 and then broken out by activity code. [A] HE applications from FY2010- FY2020. [B] Non-HE applications from FY2010- FY2020. [C] HE awards from FY2010- FY2020. [D] Non-HE awards from FY2010- FY2020.

### 3.3 Interrupted Time Series Analysis (ITSA)

ITSA is a method used for evaluating intervention studies such as public health interventions, vaccinations, traffic speed zones, policy changes, and health impacts in response to the global financial crisis (Bernal et al. 2017). Interventions measured using ITSA require a clear differentiation of pre- and post-intervention intervals in addition to sequential measures of the outcome at different times. Time since intervention can also be examined for sustained or ongoing effects of the intervention in the post-policy data. In this study, ITSA was used to retrospectively analyze individual data points for each council round to infer the impact of the guidance on health economics application rates over time both before and after the guidance release.

#### 3.3.1 Application rates

First, predicted values for the application rate of all health economics applications within each council round were plotted using the ITSA regression (see methods section and Figure A4). The slope in the graph accounts for all of the covariates whereas the table shows the coefficient trends independently for each variable (time, treatment, and time since treatment). Regression data was compared before and after guidance publication (dotted red line) (Figure 3A). There was a trending, but not significant, decrease in application rates prior to the guidance and a not significant increase in application rates following the guidance (Figure 3B). Of note, however, analysis of application rates by council round identified a seasonal pattern, with increased numbers of applications each year in the fourth (August) council round. As the August council round is when most of the solicited applications are considered by Council, we separated applications by whether they were solicited or unsolicited and analyzed each group independently.

**Figure 3.**
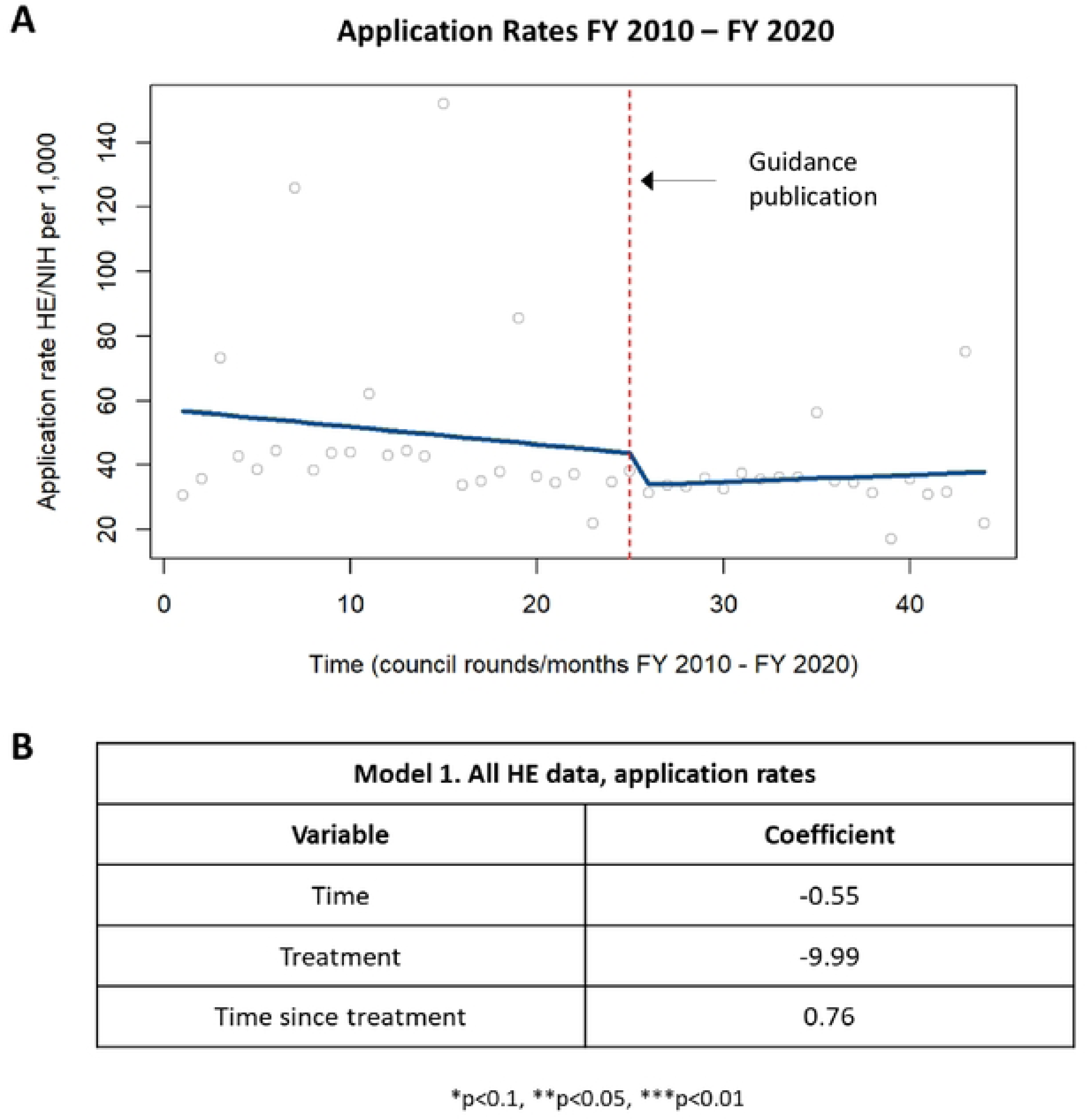
Application rates in HE applications. [A] Chart showing changes in application rates in HE applications prior to and following the guidance publication in 2015. [B] Table summarizing the coefficients from the time series analysis.

When examining the application rates of only solicited applications, a similar trend was identified in which there was a trending but not significant decrease in application rate pre-guidance and a trending but not significant increase in application rate post-guidance (Figure A5). When examining only the application rates of only unsolicited applications, there was a decrease in application rates pre-guidance that was statistically significant with no significant difference in application rates post-guidance (Figure 4).

**Figure 4.**
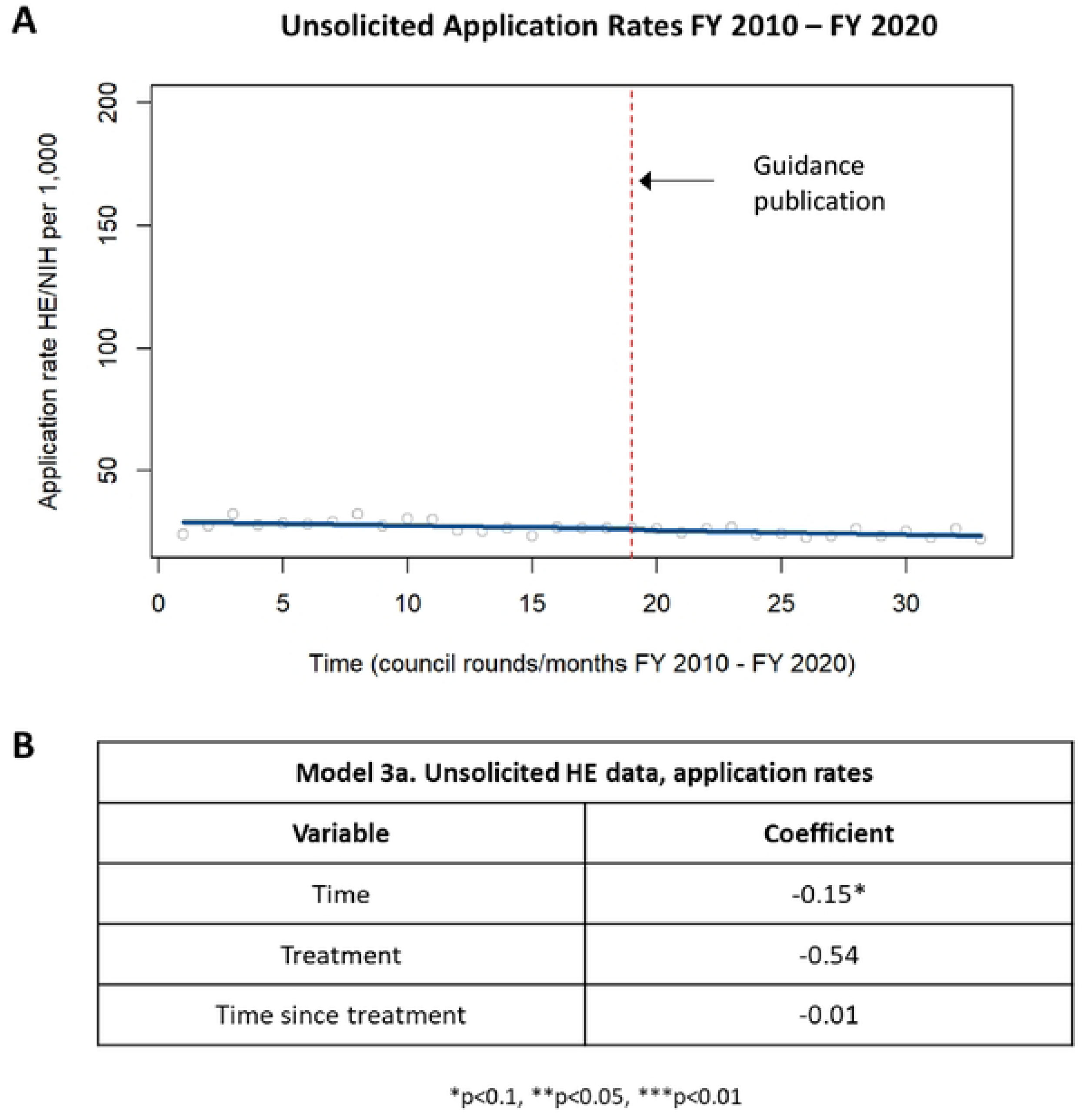
Application rates in unsolicited HE applications. [A] Chart showing changes in application rates in unsolicited HE applications prior to and following the guidance publication in 2015. [B] Table summarizing the coefficients from the time series analysis.

Since the time between the submission date of an application and the notice of grant award date can be up to a year, there was an expected lag of about a year from when the guidance was released to when it might impact application submission rates and subsequent council concurrence. To account for this delay, the application rate of solicited applications was reevaluated using ITSA in which the time designated as the “intervention” was shifted twelve months later. With this shift, the application rates during the first twelve months post-guidance were now included in the “pre-guidance” period. When the delayed effect analysis was performed, this model showed that the application rate was decreasing at a statistically significant level up until one year after publication of the guidance, with no significant change in the application rate one-year post-guidance (Figure 5).

**Figure 5.**
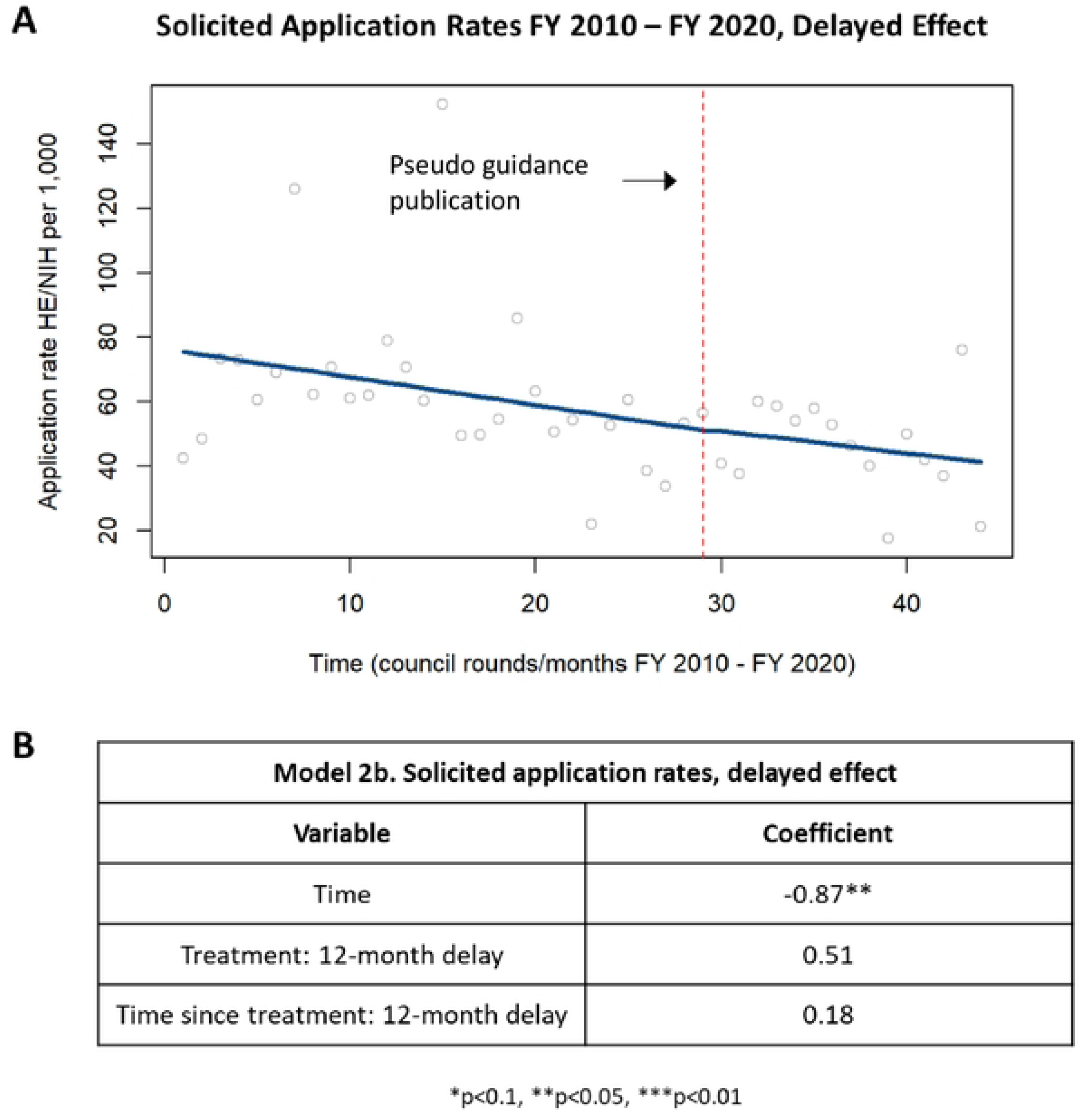
Delayed effect of application rates in solicited HE applications. [A] Chart showing changes in application rates in solicited HE applications up to one year after publication of the guidance and starting one year after publication of the guidance. [B] Table summarizing the coefficients from the time series analysis.

A delayed effect analysis of unsolicited applications demonstrated a statistically significant decrease in application rates up to one year after publication of the guidance. There was no change in application rates starting one year after publication of the guidance (Figure 6).

**Figure 6.**
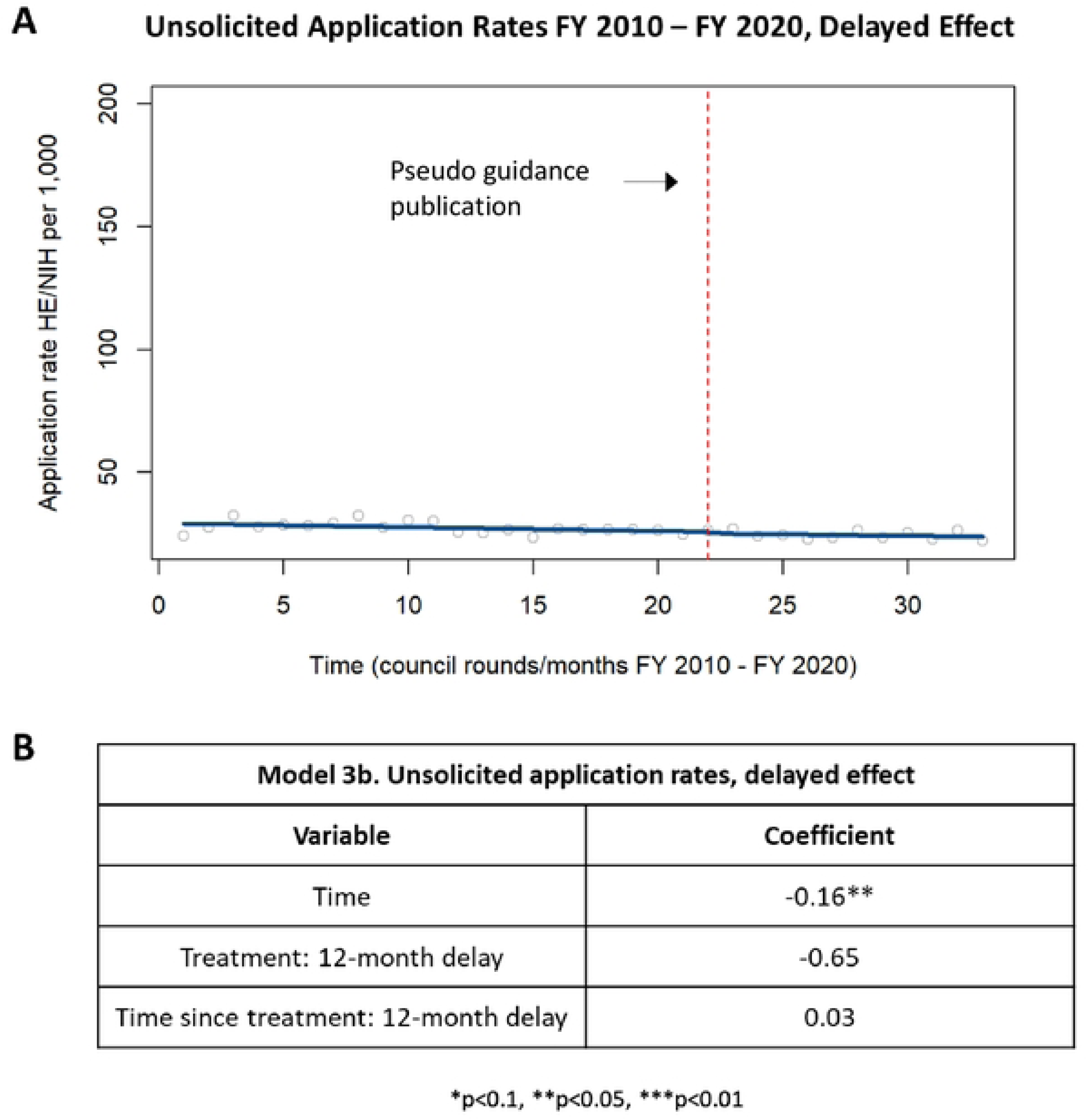
Delayed effect of application rates in unsolicited HE applications. [A] Chart showing changes in application rates in unsolicited HE applications up to one year after publication of the guidance and starting one year after publication of the guidance. [B] Table summarizing the coefficients from the time series analysis.

#### 3.3.2 Funding rates

The funding rate of health economics applications was determined for each council round by comparing the funding amounts of health economics applications to the funding amounts of non-health economics applications as described in the methods and was analyzed by ITSA. Pre-guidance, there was a decrease in health economics funding rates that was statistically significant while post-guidance funding rates demonstrated a statistically significant increase in health economics funding rates (Figure 7).

**Figure 7.**
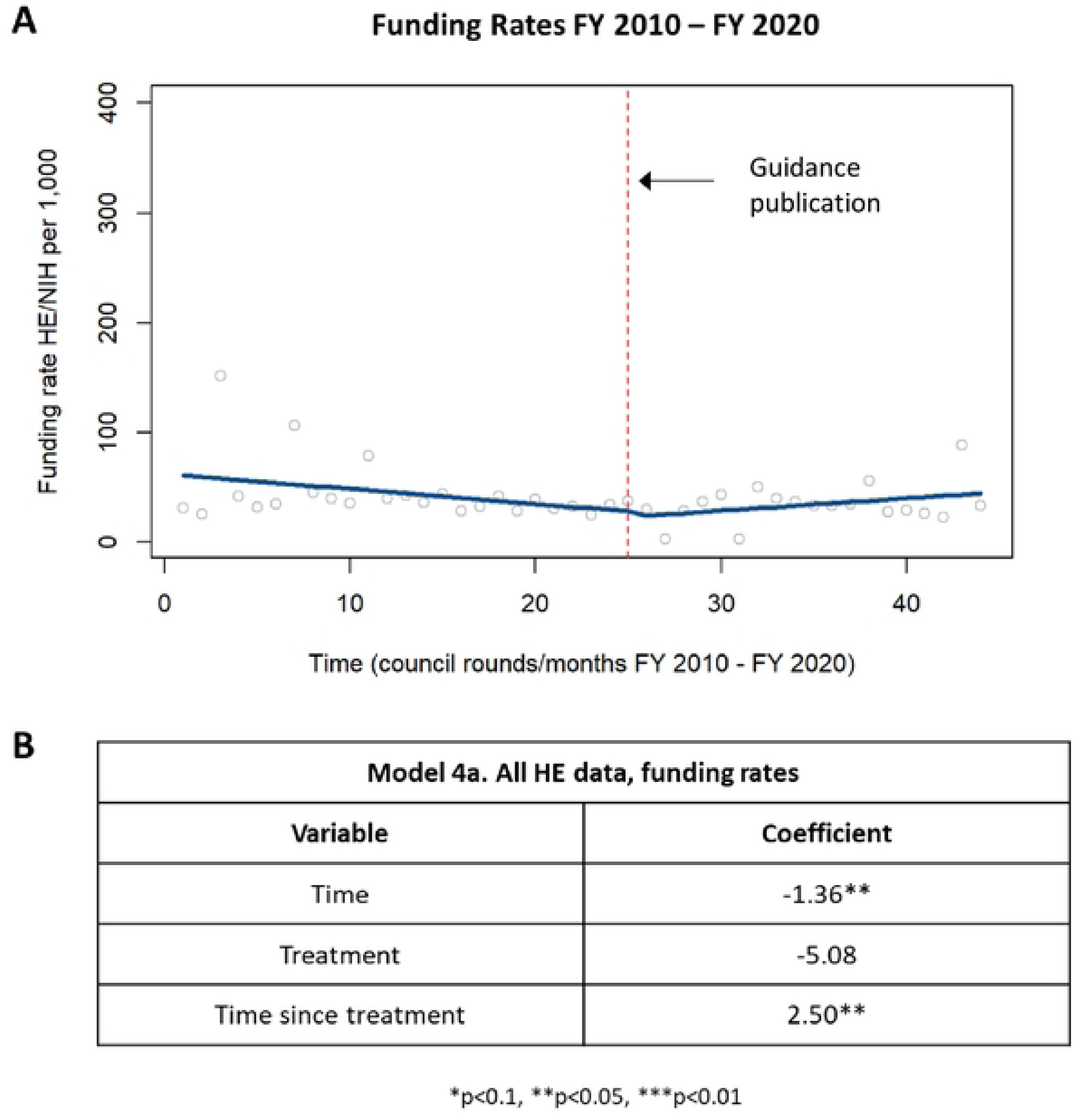
Funding rates in HE applications. [A] Chart showing changes in funding rates in HE applications prior to and following the guidance publication in 2015. [B] Table summarizing the coefficients from the time series analysis.

When the funding rate of solicited applications was analyzed, there was a decrease in funding rates for solicited applications before publication of the guidance that was statistically significant; after publication of the guidance, there was a significant increase in funding rates of solicited applications (Figure 8).

**Figure 8.**
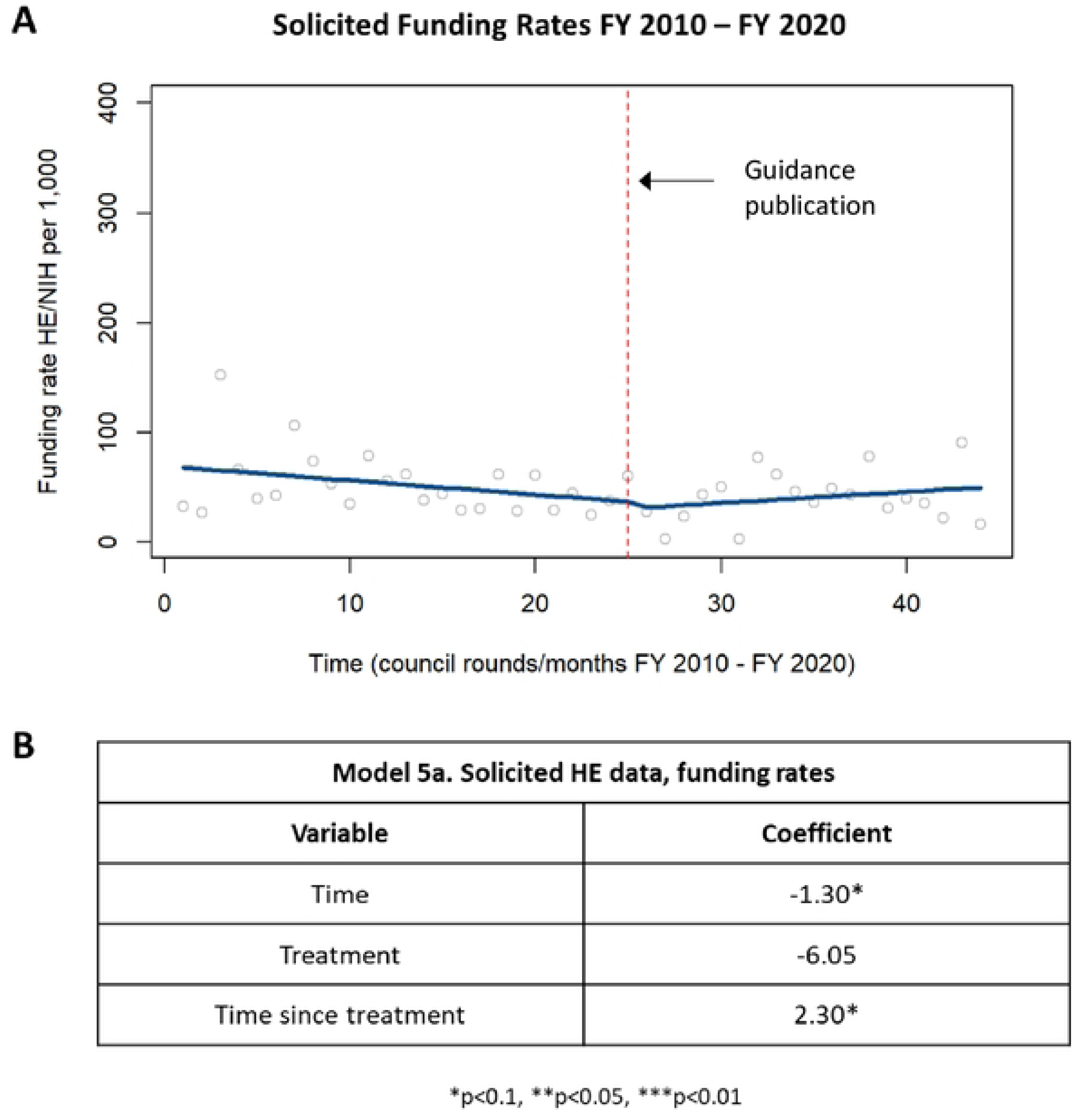
Funding rates in solicited HE applications. [A] Chart showing changes in funding rates in solicited HE applications prior to and following publication of the guidance in 2015. [B] Table summarizing the coefficients from the time series analysis.

When the funding rate of unsolicited applications was separately analyzed, there was a no difference in funding rates for unsolicited applications before or after publication of the guidance (Figure A6).

The funding rate of health economics applications was reevaluated using ITSA in which the time designated as the “intervention” was shifted twelve months later. When the delayed effect analysis was performed for all health economics applications, the funding rate was found to be decreasing, to a statistically significant level, up until one year after publication of the guidance, with no significant change in application rate one-year post-guidance (Figure 9).

**Figure 9.**
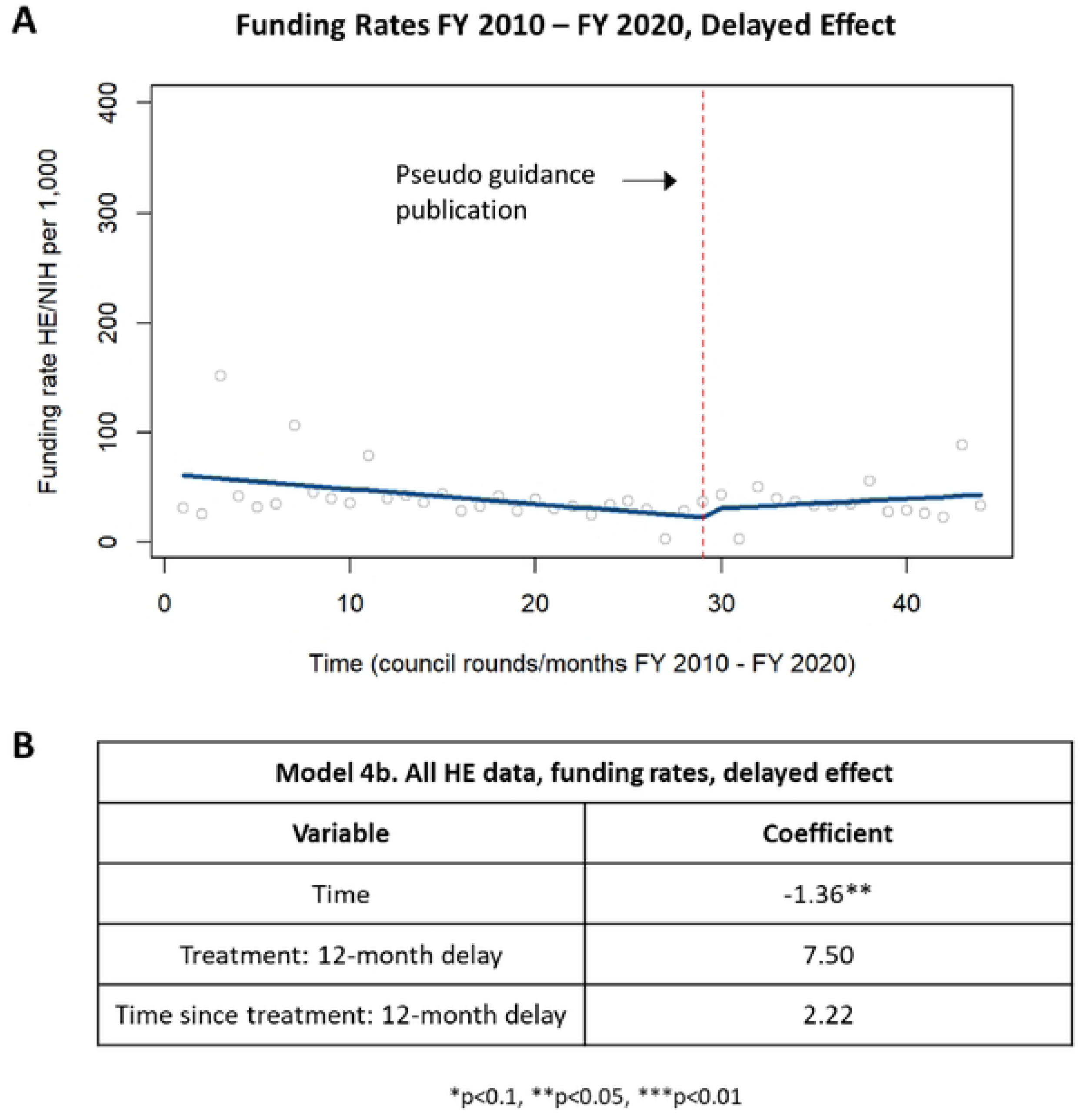
Delayed effect of funding rates in HE applications. [A] Chart showing changes in funding rates in HE applications up to one year after publication of the guidance and starting one year after publication of the guidance. [B] Table summarizing the coefficients from the time series analysis

When analyzing the delayed effect on funding rates of solicited applications, there was a decrease in funding rates up until one year following the guidance publication that was statistically significant (Figure 10). However, one year after publication of the guidance, there was a trending but not statistically significant increase in funding rates for solicited applications (Figure 10).

**Figure 10.**
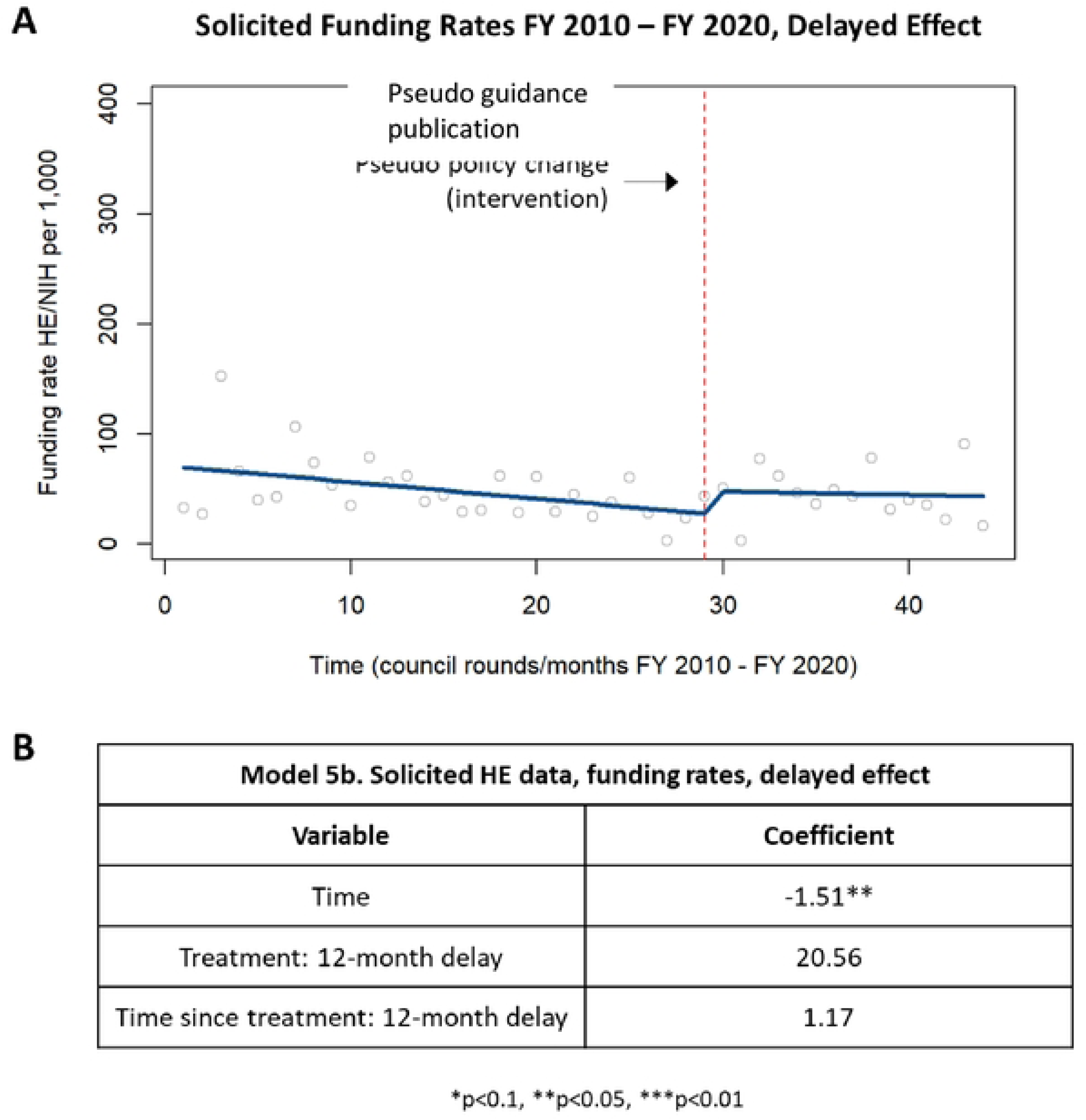
Delayed effect of funding rates in solicited HE applications. [A] Chart showing changes in funding rates in solicited HE applications up to one year after publication of the guidance and starting one year after publication of the guidance. [B] Table summarizing the coefficients from the time series analysis.

When analyzing the delayed effect on funding rates of unsolicited applications, there was again no difference in funding rates up until one year following the guidance publication or one year following the guidance publication (Figure A7).

## 4. Discussion

The results from these analyses indicate that while application rates for health economics applications were variable post-guidance, both award rates and funding amounts for health economics applications slightly increased post-guidance. Interestingly, the analysis also demonstrated that pre-guidance, both solicited and unsolicited health economics applications rates and funding amounts were slightly decreasing over time. However, in the council rounds following the guidance, the award rates and funding amounts increased. Of note, award rates for health economics awards increased post-guidance (from 14.7% to 16.9%) but were still less than the post-guidance award rates for applications not related to health economics (17.8%). This suggests that the guidance may have renewed clarity of NIH’s health economics interests for applicants, increasing the chances of applications submitted post-guidance to align with NIH priorities and subsequently to be awarded.

Health economics is a complex field of research with varying definitions based on competing priorities (Rebelo et al. 2011). For example, the CDC (Center for Disease Control and Prevention, 2013) places a strong emphasis on the use of health economics to determine alternative prevention strategies while the Johns Hopkins Bloomberg School of Public Health (Johns Hopkins Bloomberg School of Public Health, 2021) uses health economics to promote healthy lifestyle behavior. Due to the inconsistent definitions and lack of a “health economics” classification in the NIH RCDC, the current study used a multipronged approach. The use of an ML approach with input from subject matter experts allowed for a transparent and reproducible identification of health economics applications and awards to assess application and award trends over time. ML provided a consistent approach to identify the corpus of health economics applications over time and ensure that health economics research applications from FY2010 were analyzed using the same methodology as health economics research from FY2020.

It is possible that some health economics applications were not captured using this approach (false negatives) or that some research unrelated to health economics may have been captured at the expense of capturing all health economics research (false positives). This noise, however, is likely proportional across years due to the standardized ML approach. Of note, this analysis was repeated with a lower confidence threshold (0.95), and the results remained nearly identical, indicating similar findings when the labeling of a health economics application was more liberally defined.

To analyze the impact of the guidance on the application and funding rates for NIH health economics, ITSA was used as a quasi-experimental approach for inferring causality following the guidance publication. Because a controlled study of this guidance was not possible, the ITSA model was an appropriate tool for this analysis with the guidance publication acting as a defined time point (unlike most public health interventions) and the pre-guidance data acting as a clear control condition (Hudson et al. 2019). ITSA also provided further information as to application and funding trends by council round, which identified a difference in application rates between solicited and unsolicited applications and unique trends pre- and post-guidance. This methodological approach has been used in other NIH projects to categorize and analyze complex data (Villani et al. 2018) and may be a helpful tool to other researchers, especially in the health policy field, for its ability to identify the impact of a specific event on qualifying datasets in a precise and reliable manner.

### 4.1 Conclusion

Cognizant of these limitations, this study shows that the NIH health economics guidance does not appear to have negatively impacted the rate of either health economics applications or awards. Instead, the result of the guidance publication appears to have been an increase in health economics award rates and funding amounts, possibly because health economics applications are now better aligned with NIH mission and interests.

## Data Availability

All data on funded applications can obtained from the publicly-accessible RePORTER database. Data on unfunded applications cannot be shared publicly as they are protected under confidentiality.

https://reporter.nih.gov/

## 6. Captions for Supplementary Tables and Figures

**Table S1.** Competing RPG applications and awards for HE and non-HE projects before and following publication of the guidance.

| Fiscal Year | HE Not Awarded | HE Awarded | Total HE Applications |
| --- | --- | --- | --- |
| 2010 | 1,278 | 262 | 1,540 |
| 2011 | 2,044 | 350 | 2,394 |
| 2012 | 2,094 | 323 | 2,417 |
| 2013 | 2,113 | 344 | **2,457** |
| 2014 | 1,744 | 319 | 2,063 |
| 2015 | 1,758 | 303 | 2,061 |
| Totals before policy | 11,031 | 1,901 | 12,932 |
| Average before policy | 1,839 | 317 | 2,155 |
| 2016 | 1,742 | 323 | 2,065 |
| 2017 | 1,706 | 366 | 2,072 |
| 2018 | 1,784 | **414** | 2,198 |
| 2019 | 1,709 | 334 | 2,043 |
| 2020 | 1,729 | 323 | 2,052 |
| Totals after policy | 8,670 | 1,760 | 10,430 |
| Average after policy | 1,734 | 352 | 2,086 |
| Fiscal Year | Non-HE Not Awarded | Non-HE Awarded | Total Non-HE Applications |
| 2010 | 32,634 | 7,528 | 40,162 |
| 2011 | 43,685 | 8,314 | 51,999 |
| 2012 | 45,801 | 8,663 | 54,464 |
| 2013 | 43,730 | 7,845 | 51,575 |
| 2014 | 44,606 | 8,818 | 53,424 |
| 2015 | 5,925 | 9,214 | 55,139 |
| **Totals before policy** | 256,381 | 50,382 | 306,763 |
| **Average before policy** | 42,730 | 8,397 | 51,127 |
| **2016** | 47,981 | 10,040 | 58,021 |
| **2017** | 48,357 | 9,823 | 58,180 |
| **2018** | 48,217 | 10,668 | 58,885 |
| **2019** | 48,544 | 10,707 | 59,251 |
| **2020** | 48,185 | 11,029 | 59,214 |
| **Totals after policy** | 241,284 | 52,267 | 293,551 |
| **Average after policy** | 48,257 | 10,453 | 58,710 |

**Figure S1.**
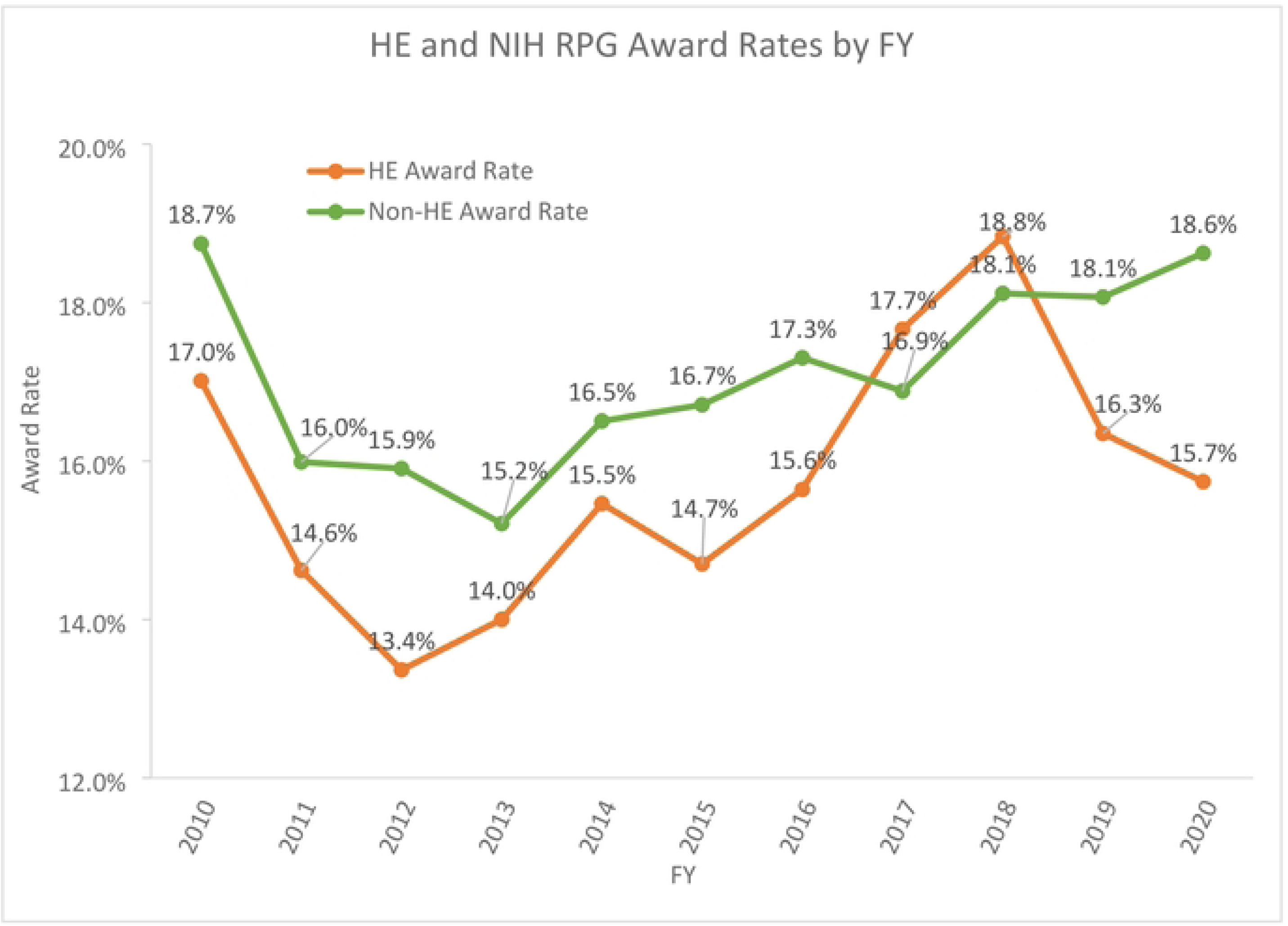
Analysis of HE and NIH (excluding HE) award rates for competing RPGs from FY2010- FY2020. Award counts were divided by the total number of applications for HE and non-HE research for each Fiscal Year.

**Figure S2.**
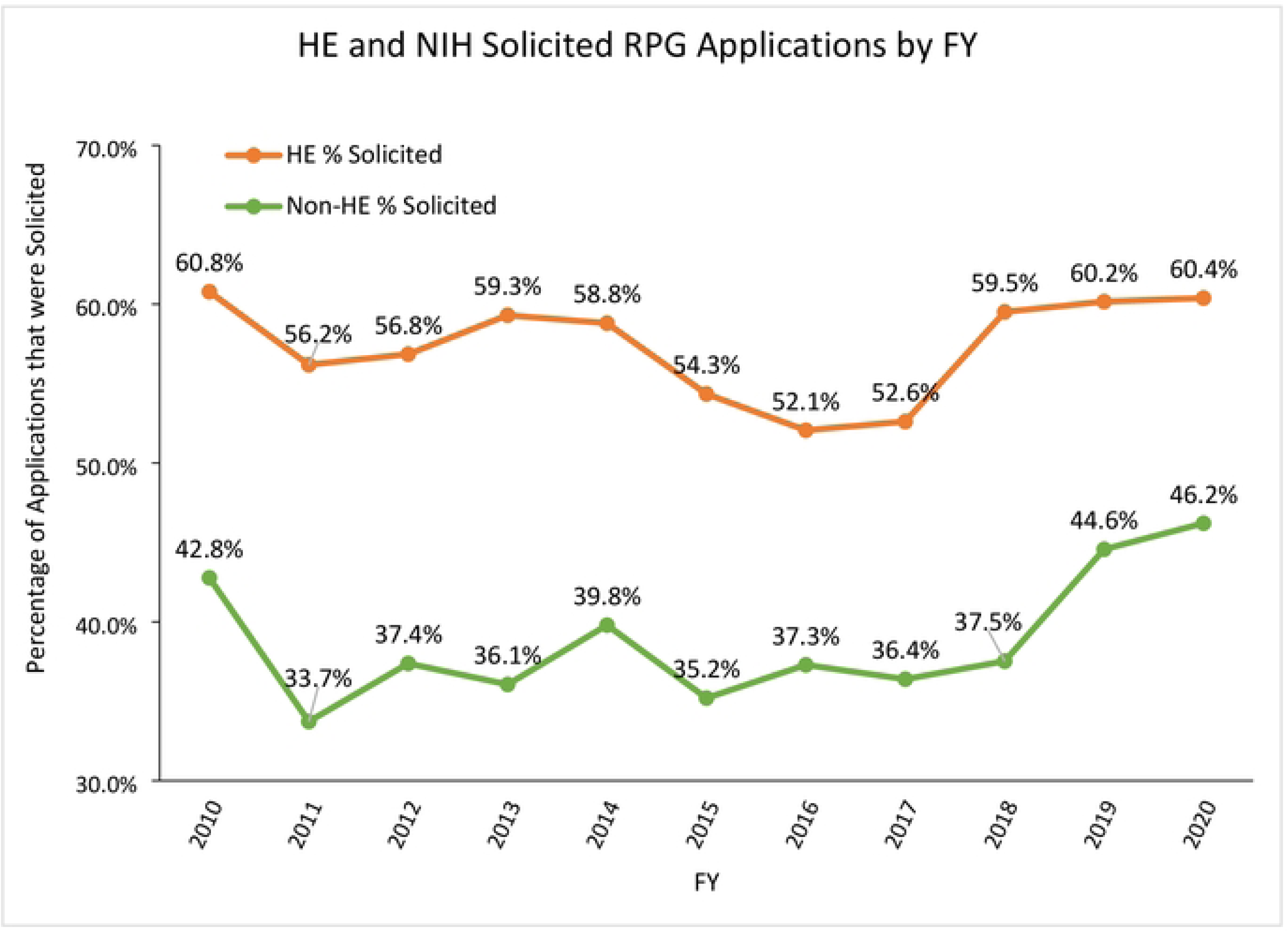
Analysis of competing applications that were solicited for HE and non-HE research from FY2010 to FY2020.

**Figure S3.**
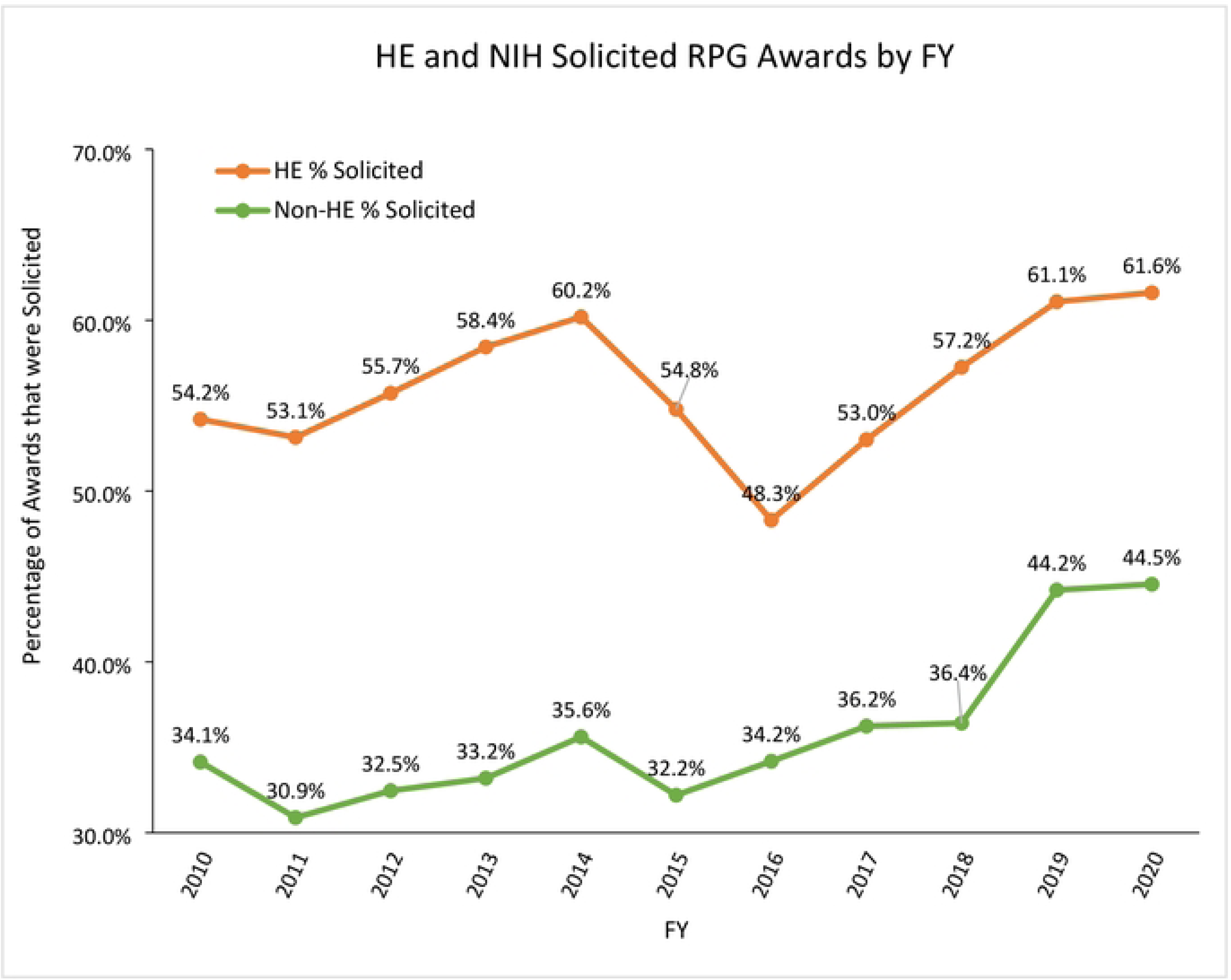
Analysis of competing awards that were solicited for HE and NIH (excluding HE) research from FY2010 to FY2020.

**Figure S4.**
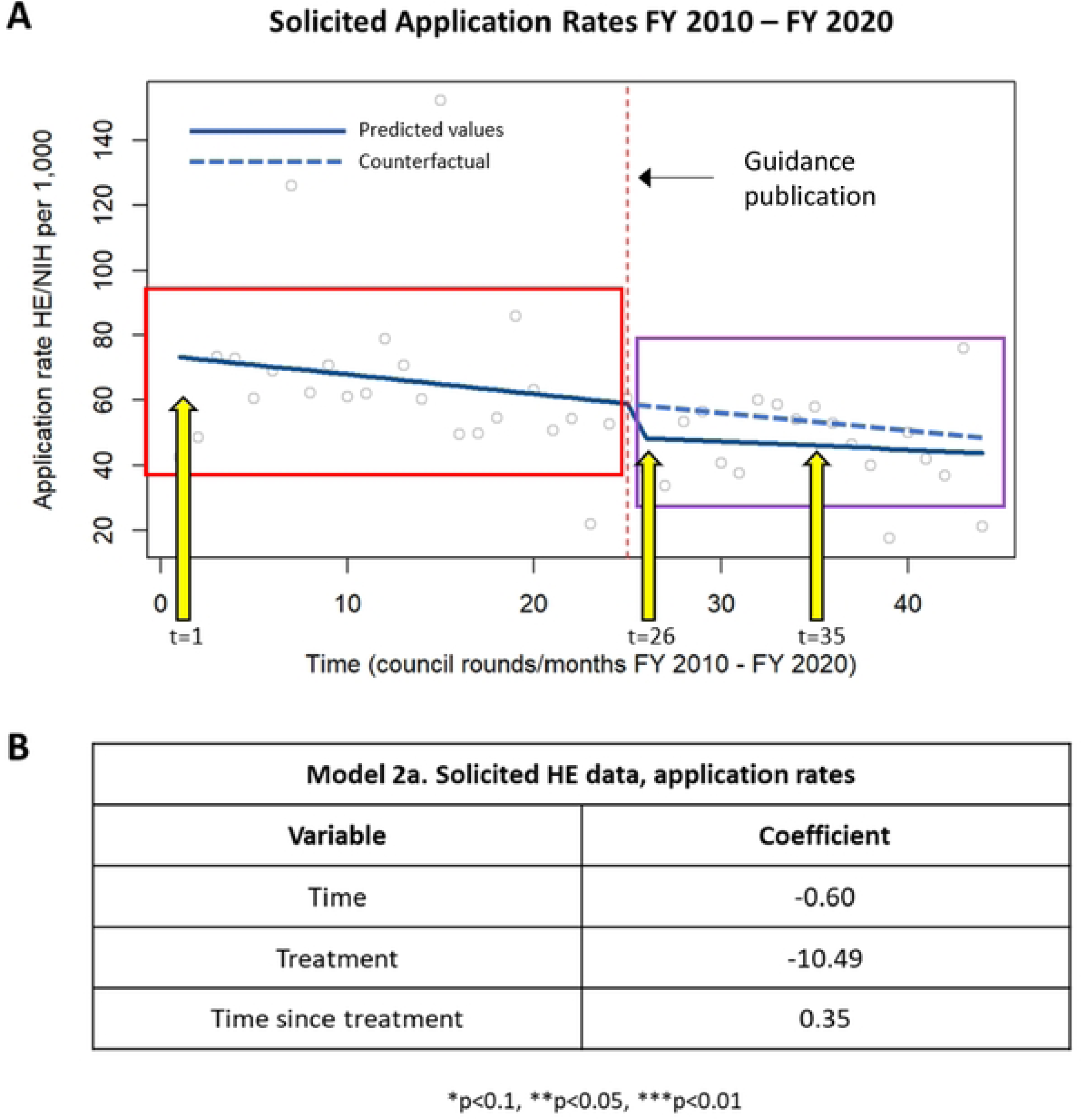
Illustrated explanation of ITSA graph with counterfactual line added. The red box outlines pre-policy time series points and the purple box, post-policy points. Arrows are pointing to time=1, 26 and 35; explanations of their predictions are below. Coefficients are otherwise known as independent slopes (per variable) within the regression model. For the part of the graph outlined above in red, these are the pre-policy time points, therefore, with regard to the regression model, only the baseline constant and time variable are utilized in deriving this point (not intervention or time since intervention). For model 2a above, solicited application rates, the predicted outcome, can be derived using the necessary coefficients and values from the regression table, for example, at time=1: Yt = β0 + β1(1) + β2(0) + β3(0) Yt = 73.90 + (-0.60)(1) + (-10.49)(0) + (0.35)(0) **Predicted Yt = 73.30** Looking at trends after the policy was put into place, outlined in the purple box on the graph above: Immediately after the policy change (May 2016), for this model (2a), time=26, intervention=1 and time since=1: Yt = β0 + β1(26) + β2(1) + β3(1) Yt = 73.90 + (-0.60)(26) + (-10.49)(1) + (0.35)(1) **Predicted Yt = 48.16** Looking further out (August 2018), post-policy change, for this model (2a), time=35, intervention=1 and time since=10: Yt = β0 + β1(35) + β2(1) + β3(10) Yt = 73.90 + (-0.60)(35) + (-10.49)(1) + (0.35)(10) **Predicted Yt = 45.91** Had the policy *not* been put into place, a predicted value (counterfactual) can be calculated. This is visualized in the graph above with the dotted blue line continuing the same pre-policy trend/slope. For the same time=35 example above: Yt = β0 + β1(35) + β2(0) + β3(0) Yt = 73.90 + (-0.60)(35) + (-10.49)(0) + (0.35)(0) **Predicted Yt = 52.90** There is a slight difference in the slopes of the before and after the policy change. Therefore, at different time points, the predicted outcome will vary and may not always be higher had the policy not been put into place.

**Figure S5.**
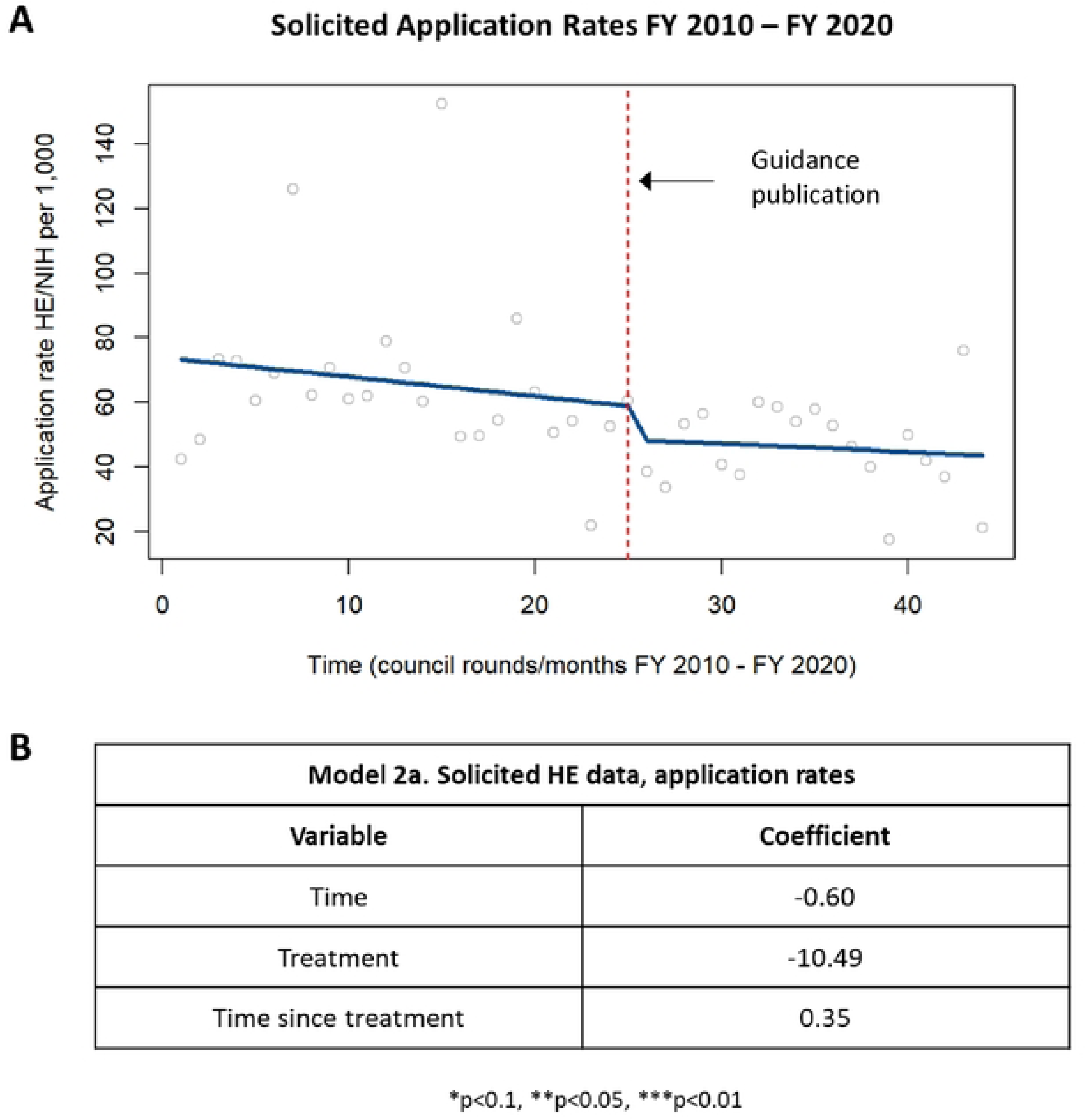
Application rates in solicited HE applications. [A] Chart showing changes in application rates in solicited HE applications prior to and following the guidance publication in 2015. [B] Table summarizing the coefficients from the time series analysis.

**Figure S6.**
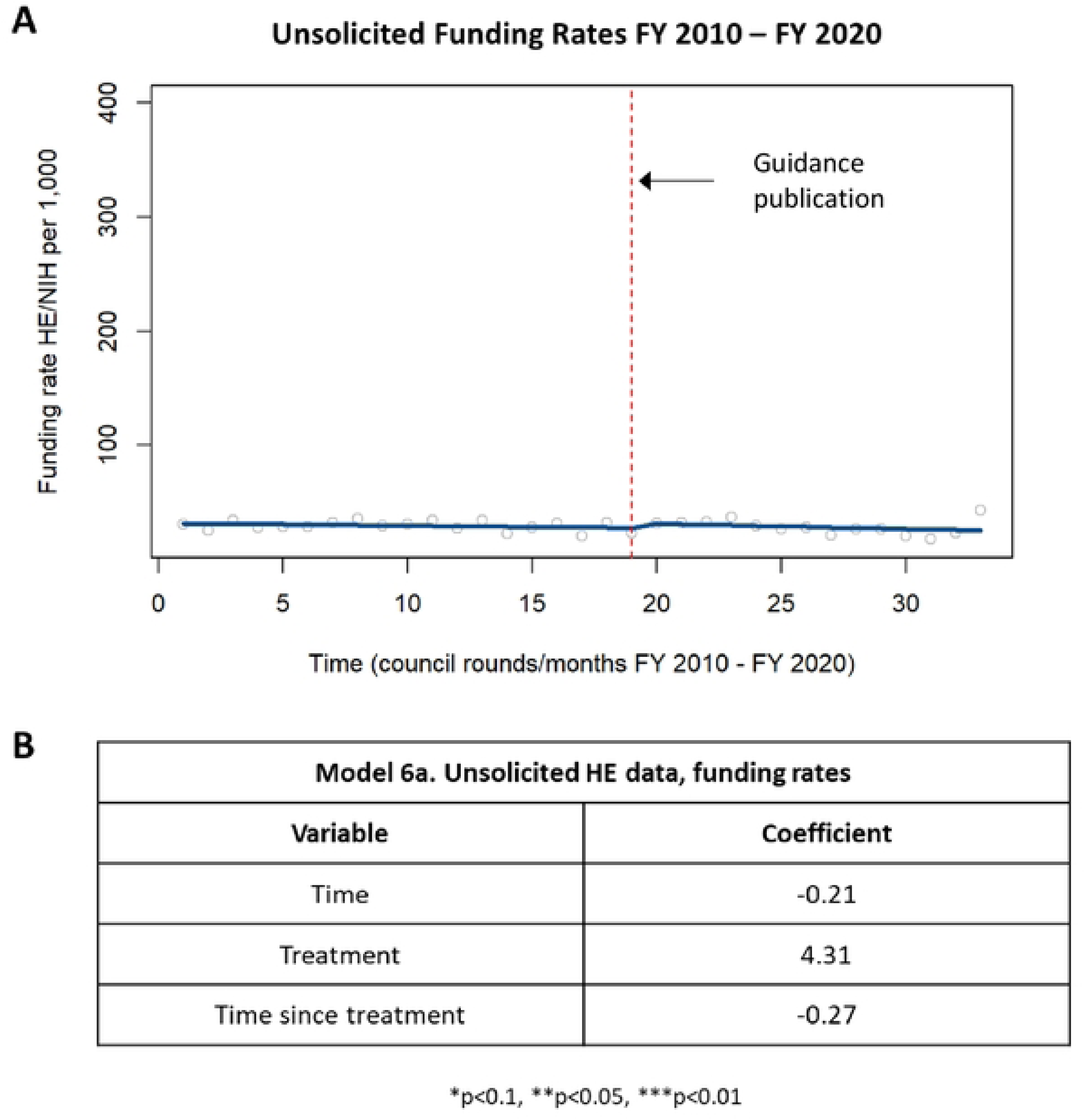
Funding rates in unsolicited HE applications. [A] Chart showing changes in funding rates in unsolicited HE applications prior to and following the guidance publication in 2015. [B] Table summarizing the coefficients from the time series analysis.

**Figure S7.**
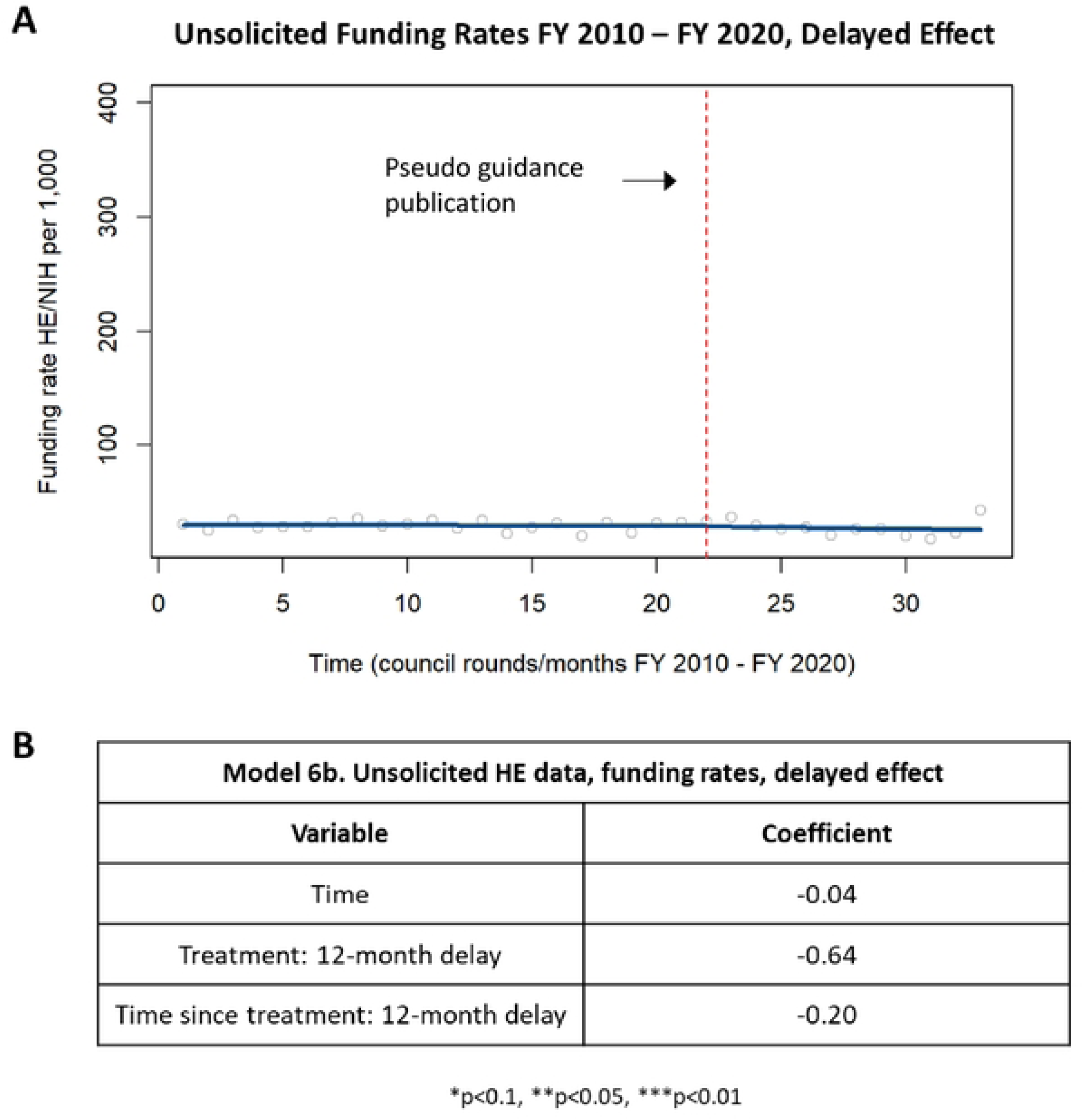
Delayed effect in unsolicited HE applications. [A] Chart showing changes in funding rates in unsolicited HE applications up to one year after publication of the guidance and starting one year after publication of the guidance. [B] Table summarizing the coefficients from the time series analysis.

